# Projection of Healthcare Demand in Germany and Switzerland Urged by Omicron Wave (January-March 2022)

**DOI:** 10.1101/2022.01.24.22269676

**Authors:** Hossein Gorji, Noé Stauffer, Ivan Lunati, Alexa Caduff, Martin Bühler, Doortje Engel, Ho Ryun Chung, Orestis Loukas, Sabine Feig, Harald Renz

## Abstract

After the implementation of broad vaccination programs, there is an urgent need to understand how the population immunity affects the dynamics of the COVID-19 pandemic in presence of the protection waning and of the emergence of new variants of concern. In the current Omicron wave that is propagating across Europe, assessing the risk of saturation of the healthcare systems is crucial for pandemic management, as it allows us to support the transition towards the endemic course of SARS-CoV-2 and implement more refined mitigation strategies that shield the most vulnerable groups and protect the healthcare systems. We investigated the current pandemic dynamics by means of compartmental models that describe the age-stratified social-mixing, and consider vaccination status, vaccine types, and their waning efficacy. Our goal is to provide insight into the plausible scenarios that are likely to be seen in Switzerland and Germany in the coming weeks and help take informed decisions. Despite the huge numbers of new positive cases, our results suggest that the current wave is unlikely to create an overwhelming health-care demand: owing to the lower hospitalization rate of the novel variant and the effectiveness of the vaccines. Our findings are robust with respect to the plausible variability of the main parameters that govern the severity and the progression of the Omicron infection. In a broader context, our framework can be applied also to future endemic scenarios, offering quantitative support for refined public health interventions in response to recurring COVID-19 waves.

## Introduction

As the Omicron variant-of-concern (B.1.1.529) surge is unfolding in Europe, it is crucial to analyze possible scenarios likely to be seen in the next couple of weeks. In the current pandemic phase, it is of great importance to investigate whether the surge in the case number would translate into a significant wave of hospitalizations, possibly threatening the healthcare system. While the current decoupling between case number and hospitalizations observed in the UK gives reasons for optimism, the evolution of the current Omicron wave might be different in countries with a smaller fraction of vaccinated or recovered people, or with a different vaccine mix. In particular, central European countries with lower vaccination rates and relatively less dramatic previous waves might turn out more vulnerable.

In this report, we use dynamic compartmental models to analyze plausible epidemiological scenarios for Switzerland and Germany. Mathematical models are crucial to explain the non-obvious concurrent large case number and relatively low hospitalization rate that are observed in the Omicron wave. In order to elucidate the mechanisms leading to the observed situation and reasonably project the evolution likely to be observed in the upcoming weeks, our analysis is refined based on the age groups and their social-mixing, as well as on vaccination status and vaccine type. This allows us to better describe, and possibly anticipate, the pressure on the healthcare system.

Our results suggest that the current wave, despite generating huge case numbers, is unlikely to create an overwhelming healthcare demand. Even with the least favorable estimate of plausible parameters, our scenarios display hospital occupancy lower than the previously experienced peaks (especially in the intensive-care-unit, ICU). However, it should be kept in mind that the sheer number of cases might lead to staff shortage, as well as to insufficient capacity of SARS-CoV-2 diagnostic resources.

Our results can be used to better refine the current measures in the pandemic management of both countries, with a focus on the healthcare demand. Furthermore, our modeling framework can also help project the endemic course of the unfolding Omicron wave.

## Results

To project the current situation into the coming weeks and investigate the consequences of the Omicron wave in terms of case number, hospitalization, and ICU bed occupancy in Switzerland and Germany, we considered three scenarios characterized by different effective reproductive numbers, i.e., 1.3, 1.5 and 1.8. These scenarios should be contrasted to the current epidemiological course with an observed reproductive numbers between 1 and 1.2 in both countries at the time of publication. For the base scenario projection, we fed the compartmental model by central estimates of all involved parameters (listed in Tables 1-8 in the *Supplementary Information*); then, we conducted a sensitivity analysis with respect to the severity and the vaccine protection (see Figures 4-11 in the *Supplementary Information*) and found that our results are robust as long as the considered parameters remain in a plausible range.

**Table 1:**
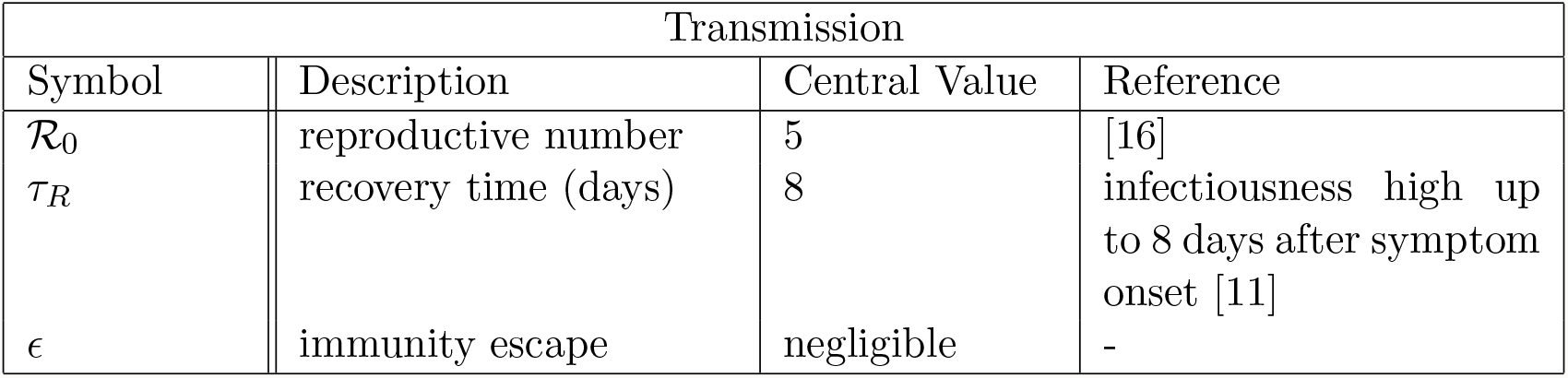
Transmission rates of Delta

The projection of daily case numbers, hospitalization and intensive care requirement are shown in Figures 1 and 2 for Switzerland and Germany, respectively. We observe a significant increase of the case number in the worst case scenario of ℛ_*e*_ = 1.8. Our projection of Germany shows 20% higher case number at the peak in comparison to Switzerland. This is mainly due to the different heterogeneity of considered contact networks among different age-groups of the two countries: in general, more heterogeneous contact patterns, as observed in Switzerland, yield markedly lower attack rates in epidemics [8]. Besides, we estimated the average protection offered by the vaccine mix against Omicron infection to be approximately 0.4 and 0.5 in Germany and Switzerland, respectively. The 20% difference in the vaccine efficacy is related to the different mix of vaccine types used in the two countries.

**Figure 1:**
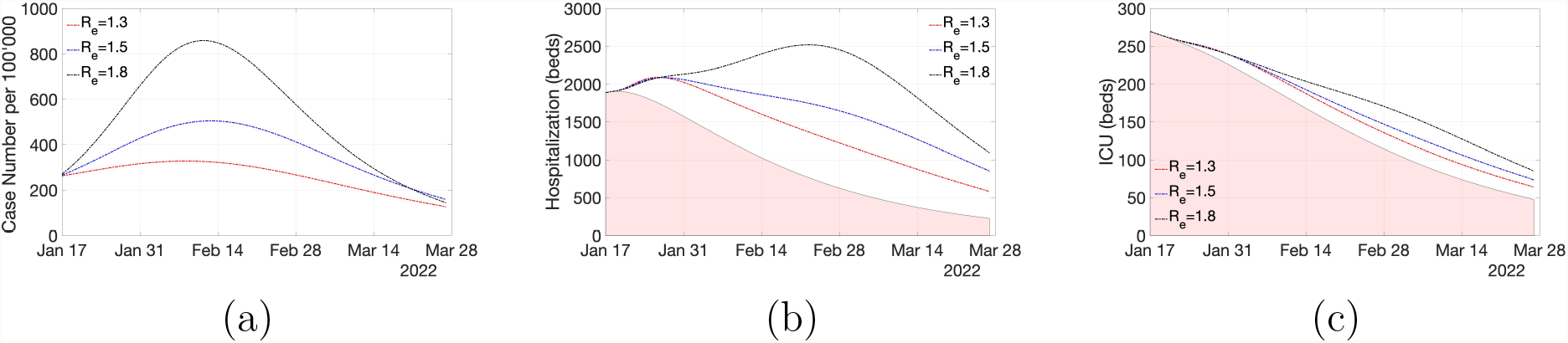
Projection of scenarios in Switzerland for (a) daily incidence (Omicron cases), (b) hospitalization (general ward) and (c) ICU occupancy. Three scenarios of ℛ_*e*_ ∈ {1.3, 1.5, 1.8} are considered for Omicron, whereas ℛ_*e*_ of 0.9 is assumed for Delta. The red shaded areas in (b) and (c) account for the occupancy due to Delta. All scenarios are initialized with an identical incidence rate. This results in an initially higher number of active cases in a scenario governed by a lower reproductive number.

**Figure 2:**
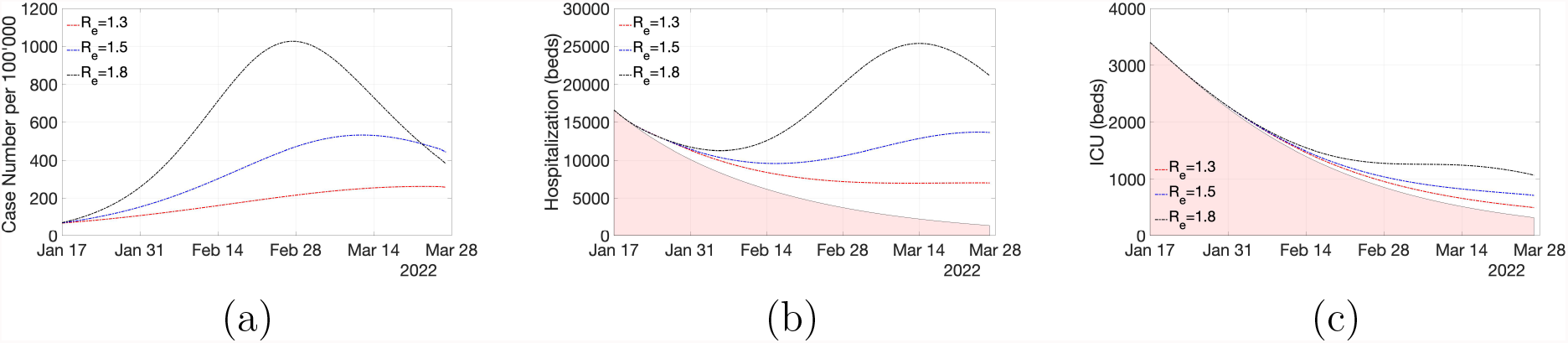
Projection of scenarios in Germany for (a) daily incidence (Omicron cases), (b) hospitalization (general ward) and (c) ICU occupancy. Three scenarios of ℛ_*e*_ ∈ {1.3, 1.5, 1.8} are considered for Omicron, whereas ℛ_*e*_ of 0.9 is assumed for Delta. The red shaded areas in (b) and (c) account for the occupancy due to Delta. All scenarios are initialized with an identical incidence rate. This results in an initially higher number of active cases in a scenario governed by a lower reproductive number.

The number of severe cases, and especially those requiring hospitalization in ICUs, remain at levels lower than the peaks observed during previous waves. This favorable outcome, despite the huge case number, is due to lower intrinsic severity of Omicron as well as to the protection from severe course offered by the vaccines (and refreshed by booster shots in recent months). We estimated the overall efficacy of the vaccine mix administered in Germany and Switzerland against hospitalization to be approximately 0.6 and 0.65, respectively.

## Discussion

The recent resurgence of COVID-19 in Europe is concurrent to the already heavy healthcare demand imposed by the most recent Delta (B.1.617.2) wave. A careful analysis of the situation is critical to help secure healthcare resources for anticipated COVID-19 patients. It is likely that the case numbers will still rise sharply as a result of the high transmissivity of the Omicron variant. At the peak of the least restrictive scenario (i.e. ℛ_*e*_ = 1.8), it is expected that 5-10% of the population would be infected in a week. While by itself this may not translate into a public health crisis, the abrupt increase in infected individuals can interrupt the presence of work forces, with significant consequences also on the healthcare system (depending on the applicable quarantine and isolation rules). Furthermore, it is likely that the testing resources would become inadequate to accommodate such a large number of infected individuals, leading to heavy underestimations of the real case numbers, even among symptomatic patients.

With regard to the healthcare systems, the simulated scenarios allow us to provide several important messages. (i) The sharp increase in the overall number of infections might also lead to a high infection rate among medical and nursing staff in hospitals and nursing homes. (ii) The demand for normal care beds for COVID-19 patients might rise within a short time period depending on the effective reproductive number. On top of patients hospitalized due to COVID-19, there will be also an increased number of hospitalized patients who are treated in the hospital for other reasons but co-infected with SARS-CoV-2. This will put an additional burden on the hospital systems in both countries. (iii) We do not expect a significant further increase in ICU patients above the current level. (iv) The sharp and rapid increase in infected cases will be followed by a likewise sharp and rapid decline in case numbers, with the exception of prolonged hospitalizations of ICU patients, which has been observed also during previous waves.

The modeled scenarios suggest that the anticipated number of infections may not lead to a significant pressure on the healthcare system, neither in Switzerland nor in Germany. This can be attributed to the intrinsically lower hospitalization rate caused by the novel variant and by the effectiveness of the vaccines in protecting from a severe course. While in Germany the vaccination rate is slightly higher, we estimate that the protection is stronger in Switzerland where a vaccine type with a longer term efficacy has been administered to the majority of the population. Our model further suggests that, as long as the reproductive number remains below 2, the ICU bed occupancy hardly reaches critical thresholds (around 400-500 beds for Switzerland and 7’000-8’000 beds for Germany).

When drawing conclusions from the simulated scenarios presented here, it is important to keep in mind that several factors may limit the validity of the modeling results. Besides intrinsic limitation of compartmental models, many of the adopted parameters are still subject to significant uncertainties. Most notably, the severity of Omicron, the protection offered by the vaccines against this variant and their waning efficacy are far from being sufficiently studied and well characterized. Here, we did not consider the possibility of long term effects of the severe/mild Omicron cases, as data is still lacking on Omicron long-Covid cases. Nevertheless, our findings are in accordance with decoupling between case number and hospitalization observed in South Africa and UK.

In interpreting this somewhat optimistic results, it should be noticed that even in the scenario assuming the least stringent measures (ℛ_*e*_ = 1.8), a certain level of mitigation measures still needs to be maintained to achieve the corresponding reduction of the infection rate (roughly 20%) with respect to the unmitigated situation. Furthermore to support the decoupling between the case number and hospitalization, it is necessary to improve the immunity of the population by expanding the vaccine uptake as well as accelerating the third dose campaigns.

Our modeling framework accounts for different age-groups and their social-mixing, vaccination status, vaccine type and protection waning. This is important as throughout the COVID-19 pandemic, a strong stratification of hospitalization has been observed with respect to the age-group. Furthermore since the vaccination rate is biased towards elderlies, it is essential that the model accounts for these heterogeneities. In a broader context, the provided framework can be applied to investigate endemic scenarios and estimate the long-term vaccination rates necessary to maintain the overall immunity of the population at risk during future outbreaks.

How well the aftermath of the current Omicron wave is representative of an endemic scenario depends on many factors, including the cross-immunity with respect to the past variants, as well as the characteristics of possibly emerging future ones. As a result, it is crucial to reassess the ongoing mitigation measures to support an exit-strategy which maintains a balance between the disease severity and consequent socio-economical tolls of the measures, in a more targeted (e.g. age-specific) manner. To address these crucial points, we need more detailed analysis on the impact of measures as well as close monitoring of the unfolding situation for some time to come.

## Supplementary Information

### Model

#### Dynamics

We consider a Susceptible-Infected-Removed (SIR) type compartmental model comprised of susceptible (*S*), infected (*I*), hospitalized in general ward (*H*), hospitalized in ICU (*IU*) and removed (*R*) populations. The progress of infection and further disease development among these compartments are illustrated in Figure 3. Each compartment is stratified per age-group denoted by superscript *i*. Furthermore they are refined for vaccinated, unvaccinated and recently recovered from a separate variant, as denoted by subscripts *v, u* and *r*, respectively. Note that as the focus here is to project the symptomatic cases and their healthcare demand, the distinguish between asymptomatic/presymptomatic and symptomatic cases is ignored. Moreover, we did not consider the mortality as it hardly has an influence on the epidemic course, which leads to a conservative estimate on the number of hospitalized and critical cases. Also despite the simplification of omitting the exposed compartment, we do not expect that the results are affected much due to the relatively short incubation period of Omicron variant (around 3 days [12]). As a result of all simplifications and uncertainty of input parameters, the projections made by the model should be considered as plausible scenarios, instead of accurate predictions of the future course of pandemic.

**Figure 3:**
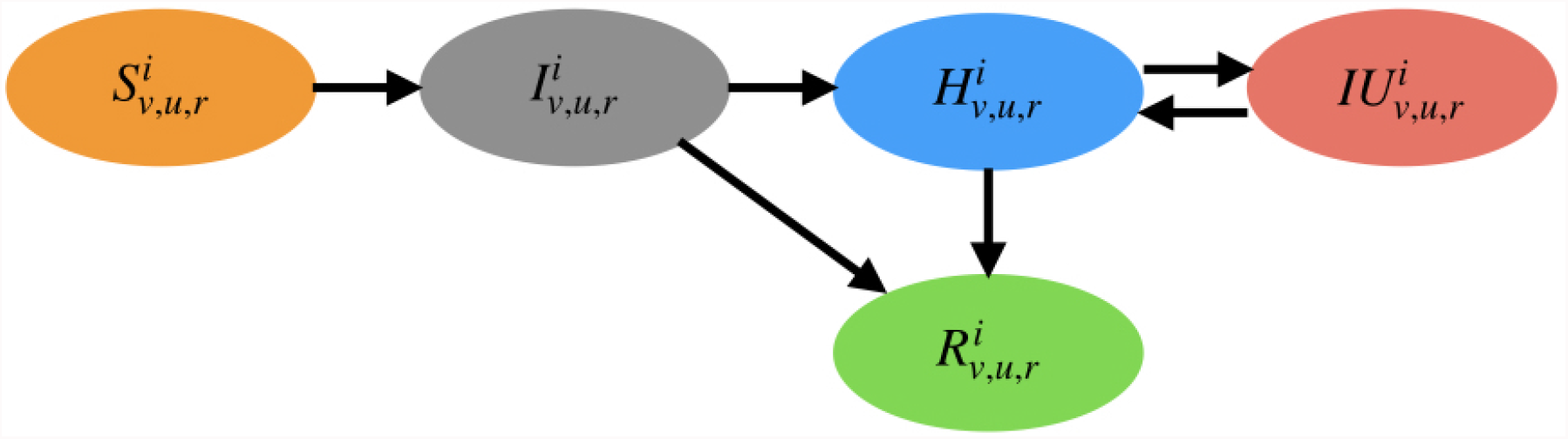
Graphical depiction of the compartmental model

The dynamic evolution of each compartment is governed by the transition rates (see e.g. [6, 4, 10] as examples of compartmental models). Let *K*_*A→B*_ be the transition rate from compartment *A* to *B*, therefore we get

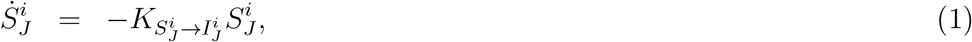

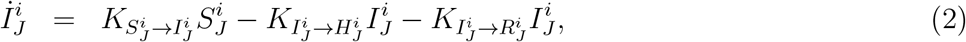

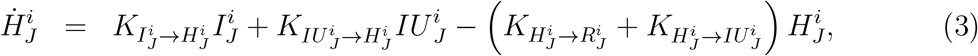

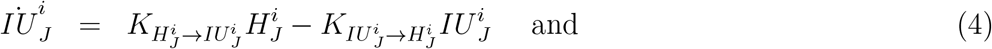

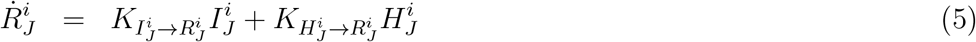

for *J* ∈ {*u, v, r*}.

The transition rates depend on intrinsic virus properties, population statistics and vaccine efficacy. Below we review the main parameters that govern the transition rates.

1. *Virus transmission and pathogenesis*: In our model, the virus transmission is governed by the infection rate *β*, immunity escape *ϵ* and recovery rate *γ* = *𝒫*_*R*_*/τ*_*R*_ where *𝒫*_*R*_ is the recovery probability and *τ*_*R*_ the average recovery time. For severe cases, the disease progression follows the hospitalization rate *ξ* = *𝒫*_*H*_ */τ*_*H*_, where *𝒫*_*H*_ = 1 − *𝒫*_*R*_ is the probability of hospitalization and *τ*_*H*_ the average time that it takes from infectiousness to hospitalization. In critical cases the admission to ICU leads to the ICU rate *µ* = *𝒫*_*IU*_ */τ*_*IU*_, where *𝒫*_*IU*_ is the probability of ICU admission (conditional on hospitalization) and *τ*_*IU*_ the average time that it takes from hospitalization to the ICU admission. Patients recovered from hospitals reduce the hospitalization by the hospital recovery rate *γ*_*H*_ = (1 − *𝒫*_*IU*_)*/τ*_*R*|*H*_ with *τ*_*R*|*H*_ the average recovery time for hospitalized cases. Finally critical cases would be back to the general ward by the ICU recovery rate *γ*_*IU*_ = 1*/τ*_*H*|*IU*_, where *τ*_*H*|*IU*_ is the average time spent in ICU.

2. *Population statistics*: The number of people *N* ^*i*^ in each age-group *i*, besides the contact matrix *𝒞*_*ij*_ (average number of people in age-group *j* in contact with a person in age-group *i*) gives the social-mixing stratified by age-groups. Note that the contact matrix follows the reciprocity constraint

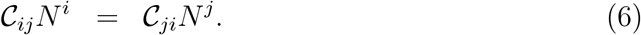

To deduce *𝒞* from contact survey *𝒟*, we apply

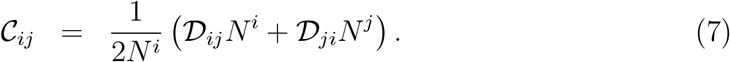

Furthermore 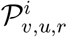 accounting for fraction of people vaccinated, unvaccinated and recently recovered from another variant, respectively, give us the effective immunity of the population per each age-group.

3. *Vaccine efficacy*: The effectiveness of the vaccines against infection 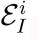, against hospitalization (conditional on infection) 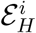 and against ICU admission (conditional on hospitalization) 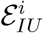 characterize the impact of vaccination in each age-groups on the pandemic course.

#### Closure

The transition rates are closed based on the above described parameters. The rate of new infections in age-group *i* is proportional to their contacts with all age-groups *j* and the corresponding prevalence in *j* (see e.g. [9]). Therefore for new infections we get

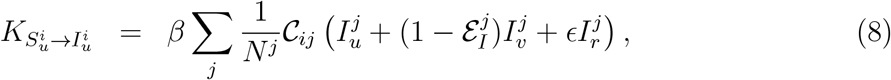

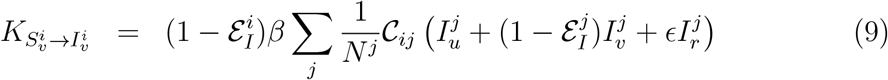

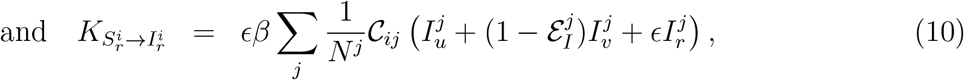

where the protection of vaccine is modeled in a symmetric fashion: it reduces both infectiousness and the probability of getting infected by the same efficacy.

Later developments in the case of mild infections follow

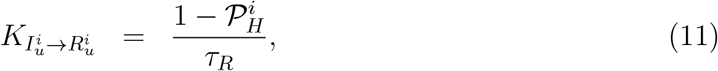

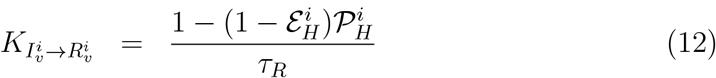

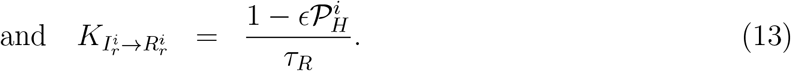

For more severe cases the rates follow

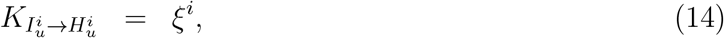

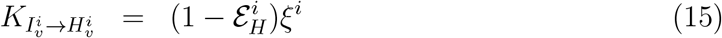

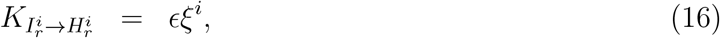

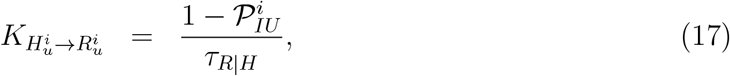

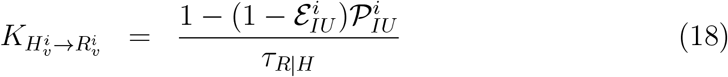

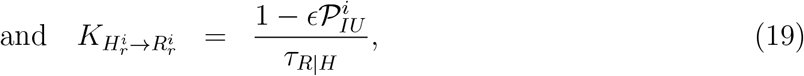

whereas the critical cases are governed by

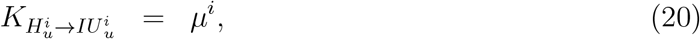

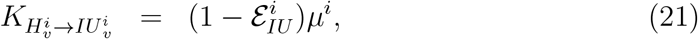

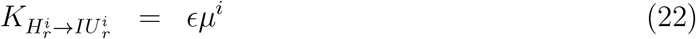

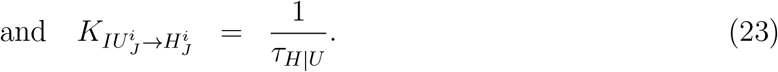

Note that for recovery rates among hospitalized cases (including critical ones), we made a conservative assumption that there is no difference between unvaccinated, vaccinated and recently recovered (from separate variant) patients concerning the average time spent in the hospital (including critical ones). Moreover, we made another conservative assumption that the disease severity of the reinfection cases is proportional to the immunity escape, similar to the reinfection rate.

### Reproductive Numbers

The reproductive number can be found based on the method of next-generation matrix. To simplify the calculations and since overwhelming majority of cases would recover without hospitalization, we ignore the severe cases in our estimate of the reproducive number. Suppose

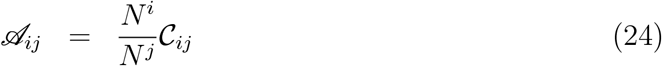

the reproductive number for our system then reads

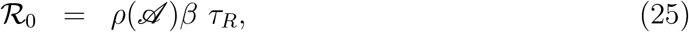

where *ρ*() returns the largest eigenvalue (assuming complete susceptible population, in the absence of vaccinated and recovered ones).

The reproductive number can be reduced by vaccination, recovery and mitigation measures. Let *κ* ∈ [0, 1] be the intensity of measures applied uniformly to all *n* age-groups (e.g. reducing number of contacts, face masks, home-office and school closures). Let us define the modified version of *𝒜* as a normalized next generation matrix subject to the introduced measures

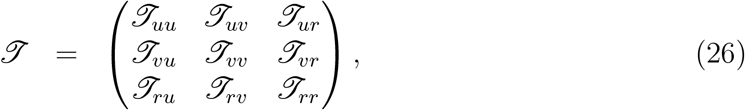

where the subscript *IJ* accounts for infections exerted by the subgroup *J* in the subgroup The block matrices *ℐ*_*IJ*_ can be found from the rate of new infections

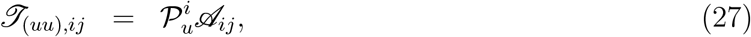

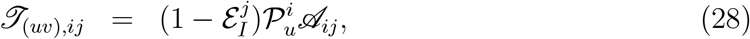

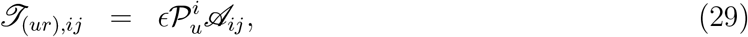

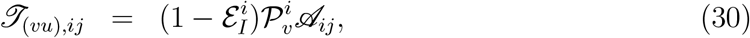

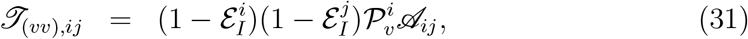

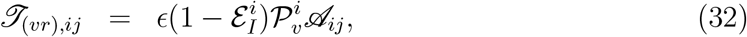

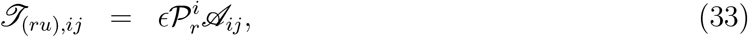

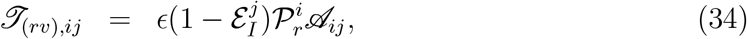

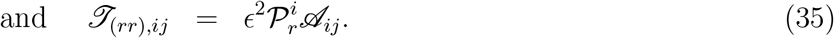

The matrix *ℐ* encodes the reduction of ℛ due to vaccination and recovery, which together with mitigation measures results in

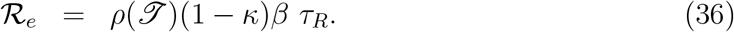

### Parameters

#### Central Rates

The adopted rates of each variant governing its transmission, disease progression and healthcare demand are discussed below:

1. *Transmission of Delta*: The basic reproduction number is estimated around 5 [16]. We take the average infectiousness period (recovery time) to be approximately 8 days, as high infectivity is detected 8 days after symptom onset [11]. The immunity escape of Delta with respect to Omicron is still unknown and it is absent in our analysis.

2. *Disease progression and healthcare demand of Delta*: We consider the disease progression of Delta to be approximately similar to the ancestral strain, but with slightly worse severity. The time window from symptom onset to hospitalization is estimated to be 5 days in average [23]. In the case of ICU requirement, we assume the transfer happens 7 days after hospitalization, consistent with the average symptom onset to ICU admission estimate of 12 days [27].

The average recovery time of hospitalized cases is taken to be 14 days (11 days estimated in [27] for the wild type). Further in more severe cases, we assume that the ICU treatment takes in average 16 days. The adopted hospitalization and ICU admission probabilities are based on [23, 7, 14].

3. *Transmission of Omicron*: The Omicron variant is associated with high increase in the case numbers. However it is still unclear how much of its growth rate is due to the immunity escape and how much due to intrinsic transmissibility. Suppose the infection rate of Omicron is by a factor 1 + *η* larger than Delta. Barnard et al. [5] suggest *η* ∈ [−0.1, 0.35]. To estimate the immunity escape *ϵ* for the suggested range of *η*, let us consider two-variant SIR dynamics [3]

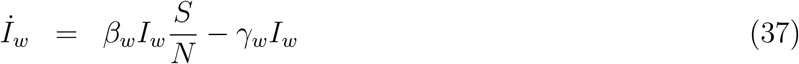

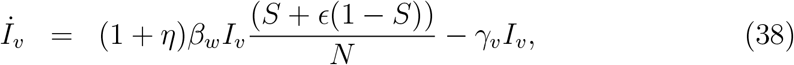

where subscript *w* and *v* represent the quantities corresponding to Delta and Omicron, respectively. For this system, the net growth rate advantage of Omicron becomes

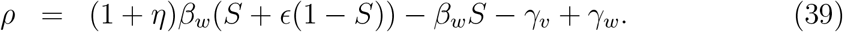

Considering that the growth rate of Omicron has been around 0.24 per day in Gauteng [24] with 73% immunity [17], we get

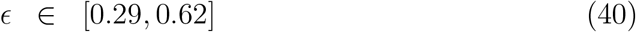

for a recovery time similar to Delta (it is anticipated that the infectiousness period is not shorter than Delta [18]). Adopting the central estimates, we have *η* = 0.125 and *ϵ* = 0.43 for our base scenario (the immunity escape is comparable to the range provided in [23] for 80% immune population). Note that this increase of infection rate results in ℛ_0_ = 5.62 for Omicron.

4. *Disease progression and healthcare demand of Omicron*: It is generally accepted that the Omicron variant less often causes severe disease progression. This accounts for both lower hospitalization rate as well as shorter treatments.

We consider recovery time of hospitalized cases to be 1.5 days (central estimate in [15]). Various reduction of hospitalization rates and ICU requirement have been estimated [26]. Furthermore [2] suggests average ICU treatment of 4 days. Following [25], we consider 56% reduction in hospitalization probability and 67% of ICU admission across all age-groups. Less optimistic reduction rates are explored in the sensitivity study.

The adopted estimations are summarized in Tables 1-2 for Delta and Tables 3-4 for Omicron. Note that while each parameter bears significant uncertainty, we only worked with central value estimates for base scenario projections. Sensitivities due to variation of a few critical rates are explored in the following section.

**Table 2:**
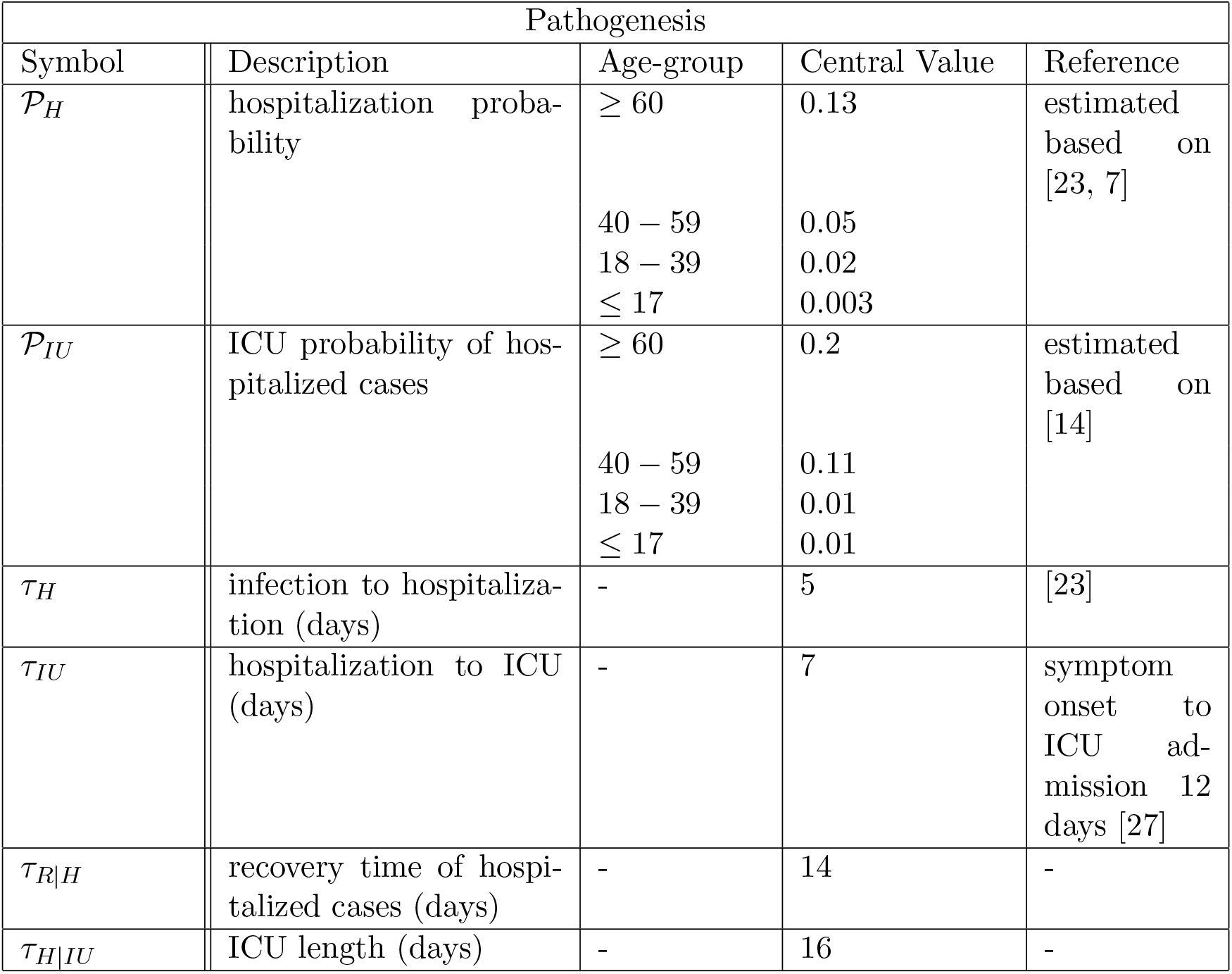
Disease progression and severity of Delta

| Pathogenesis |  |  |  |  |
| --- | --- | --- | --- | --- |
| Symbol | Description | Age-group | Central Value | Reference |
| $\mathcal{P}_H$ | hospitalization probability | $\geq 60$ | 0.13 | estimated based on [23, 7] |
|  |  | 40 – 59 | 0.05 |  |
|  |  | 18 – 39 | 0.02 |  |
| | | $\leq 17$ | 0.003 | |
| $\mathcal{P}_{IU}$ | ICU probability of hospitalized cases | $\geq 60$ | 0.2 | estimated based on [14] |
|  |  | 40 – 59 | 0.11 |  |
|  |  | 18 – 39 | 0.01 |  |
| | | $\leq 17$ | 0.01 | |
| $\tau_H$ | infection to hospitalization (days) | - | 5 | [23] |
| $\tau_{IU}$ | hospitalization to ICU (days) | - | 7 | symptom onset to ICU admission 12 days [27] |
| $\tau_{R\vert H}$ | recovery time of hospitalized cases (days) | - | 14 | - |
| $\tau_{H\vert IU}$ | ICU length (days) | - | 16 | - |

**Table 3:**
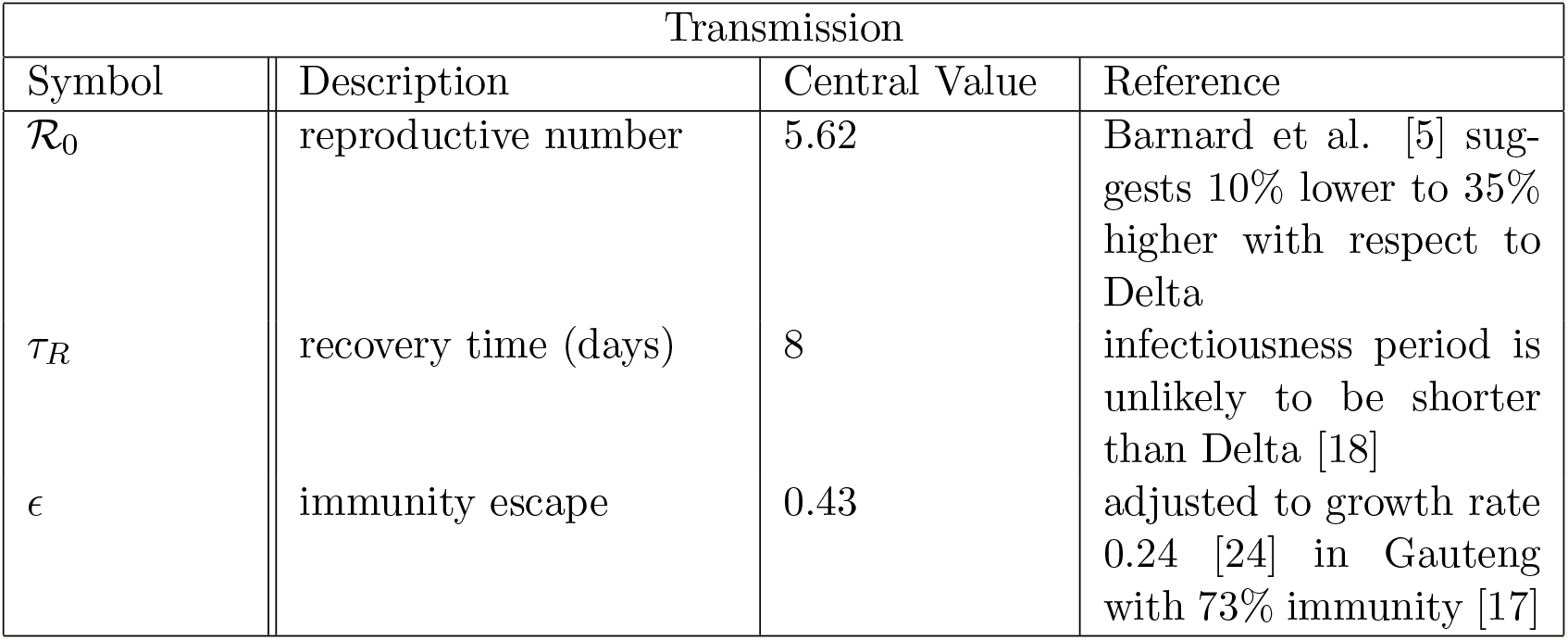
Transmission rates of Omicron

| Transmission |  |  |  |
| --- | --- | --- | --- |
| Symbol | Description | Central Value | Reference |
| $\mathcal{R}_0$ | reproductive number | 5.62 | Barnard et al. [5] suggests 10% lower to 35% higher with respect to Delta |
| $\tau_R$ | recovery time (days) | 8 | infectiousness period is unlikely to be shorter than Delta [18] |
| $\epsilon$ | immunity escape | 0.43 | adjusted to growth rate 0.24 [24] in Gauteng with 73% immunity [17] |

**Table 4:** Disease progression and severity of Omicron

| Pathogenesis |  |  |  |  |
| --- | --- | --- | --- | --- |
| Symbol | Description | Age-group | Central Value | Reference |
| $\mathcal{P}_H$ | hospitalization probability | $\geq 60$ | 0.06 | 56% lower risk of hospitalization compared to Delta [25] |
|  |  | 40 – 59 | 0.02 |  |
|  |  | 18 – 39 | 0.01 |  |
| | | $\leq 17$ | 0.001 | |
| $\mathcal{P}_{IU}$ | ICU probability of hospitalized cases | $\geq 60$ | 0.07 | 67% lower risk of ICU admission compared to Delta [25] |
|  |  | 40 – 59 | 0.04 |  |
|  |  | 18 – 39 | 0.003 |  |
| | | $\leq 17$ | 0.003 | |
| $\tau_H$ | infection to hospitalization (days) | - | 5 | similar to Delta (assumption) |
| $\tau_{IU}$ | hospitalization to ICU (days) | - | 5 | similar to Delta (assumption) |
| $\tau_{R\vert H}$ | recovery time of hospitalized cases (days) | - | 1.5 | [15] |
| $\tau_{H\vert IU}$ | ICU length (days) | - | 4 | [2] |

## Vaccine Efficacy

High effectiveness of vaccines against infection and disease progression of COVID-19 has been clearly demonstrated. However the waning of antibodies, increase of breakthrough infections and overall decay of the protection offered by a mix of vaccines in a given population should be taken into account in scenario modelling. We adjust the vaccine effectiveness for a given age-group, according to the temporal decay of the protection as well as the mix of the administered vaccine types. Several modeling assumptions are taken:

1. We neglected the protections gained by the first vaccine dose.

2. Since the vaccination datasets are aggregated, it is not possible to link different administered doses to a person. Therefore to model the efficacy of *n* booster shots, we removed *n* earliest administered second shots.

3. By taking a (likely) conservative assumption, we consider the efficacy of the booster to be equivalent to a refreshed second shot.

Efficacy of each administered second/third dose is estimated as the function of time passed since vaccination (using an exponential fit) and the vaccine type. The average of all efficacies in a given age-group then gives us the vaccine efficacy for that agegroup. Tables 5-8 provide the efficacies and decays used in our estimates. Note that we neglected the reduction of the ICU admission rate (conditional on hospitalization) due to the vaccination, as still reliable estimates are not available for such efficacy over time, especially for Omicron cases.

**Table 5:** Vaccine efficacy against infection (Delta)

| Vaccine Type | Time Passed<br>(days) | Central Value | Reference |
| --- | --- | --- | --- |
| BNT162b2 | 15-30 | 0.92 | [19] |
|  | 61-120 | 0.85 |  |
|  | 181-210 | 0.29 |  |
| mRNA-1273 | 15-30 | 0.96 | [19] |
|  | 61-120 | 0.85 |  |
|  | 181-210 | 0.71 |  |
| ChAdOx1<br>nCov-19 | 15-30 | 0.68 | [19] |
|  | 61-120 | 0.41 |  |
|  | 181-210 | negligible |  |

**Table 6:** Vaccine efficacy against hospitalization caused by Delta (all types combined)

| Vaccine efficacy against hospitalization (all types combined) |  |  |
| --- | --- | --- |
| Time Passed<br>(days) | Central Value | Reference |
| 15-30 days | 0.89 | [19] |
| 61-120 days | 0.9 |  |
| >180 days | 0.42 |  |

**Table 7:** Vaccine efficacy against infection (Omicron)

| Vaccine Type | Time Passed | Central Value | Reference |
| --- | --- | --- | --- |
| BNT162b2 | 0 months | 0.63 | [1] |
|  | 6 months | 0.28 | [13] |
| mRNA-1273 | 0 months | 0.68 | [1] |
|  | 6 months | 0.4 | [13] |
| ChAdOx1<br>nCov-19 | 2 months | 0.25 | [1] |
|  | 6 months | negligible | - |

**Table 8:** Vaccine efficacy against hospitalization caused by Omicron (all types combined)

| Time Passed | Central Value | Reference |
| --- | --- | --- |
| 2-24 weeks | 0.72 | [1] |
| 24+ weeks | 0.52 | [1] |

## Simulation Details

Prior to run the simulations, the inputs on the virus transmission, the disease progression and severity, the population statistics and vaccine efficacies are set. Based on the refinement level of the mixing-patterns, 16 age-groups of 1-5, 6-10,…,75+ are considered for Switzerland and Germany, using datasets provided in [21]. The reciprocity condition is enforced via Eq. (7). Next, the infection rate is estimated for each variant from Eq. (25), and the mitigation intensity is computed using Eq. (36). We find *κ* ∈ {43%, 34%, 21%} for base scenarios ℛ_*e*_ ∈ {1.3, 1.5, 1.8} (identical for both Switzerland and Germany).

Once the transition rates are fixed, the initial condition on the number of infections in each age-group is inferred to reproduce the given daily incidence rate at the initial date. The initial shares of Omicron and Delta variants are assumed to be 90% and 10%, respectively. We supposed initially 30% of unvaccinated population has been recovered from Delta and 10% from Omicron. Afterwards, the differential equations (1)-(5) are advanced for the considered time interval, and per each variant. Finally to include the incubation period in the hospitalization and ICU admission data, similar to [22], the obtained hospitalization and ICU admission time series are convoluted with the incubation period distribution (log-normal with log mean value 1.8 and standard deviation 0.53 [20]).

## Sensitivity

We constructed four scenarios to assess robustness of our main results. Since the transmission is principally governed by the imposed effective reproductive number, here we focus on the assumptions behind the disease severity and the vaccine efficacy, and consider four scenarios:

- *Scenario 1:* The average hospital treatment of Omicron cases would take 3 times longer (i.e. 4.5 days) than our base scenario (i.e. 1.5 days).
- *Scenario 2:* The hospitalization and ICU admission rates of Omicron infections are 50% lower than the Delta ones.
- *Scenario 3:* The hospitalization and ICU admission rates of Omicron infections are 40% lower than the Delta ones. Moreover the vaccine efficacy against hospitalization of Omicron cases is 5% lower than the base scenario (i.e. 5% lower than values given in Table 8).
- *Scenario 4:* The hospitalization and ICU admission rates of Omicron infections are 30% lower than the Delta ones. Moreover the vaccine efficacy against hospitalization of Omicron cases is 5% lower than the base scenario, besides 10% lower efficacy against infection.

Figures 4-11 show the results of these four sensitivity scenarios for Switzerland and Germany. Despite the unlikely assumption of ℛ_*e*_ = 1.8, the considered scenarios are well in the range of the ICU occupancy limit in the two countries. The pressure on the general ward of hospitals can become significant though, especially in scenarios 1 and 4.

**Figure 4:**
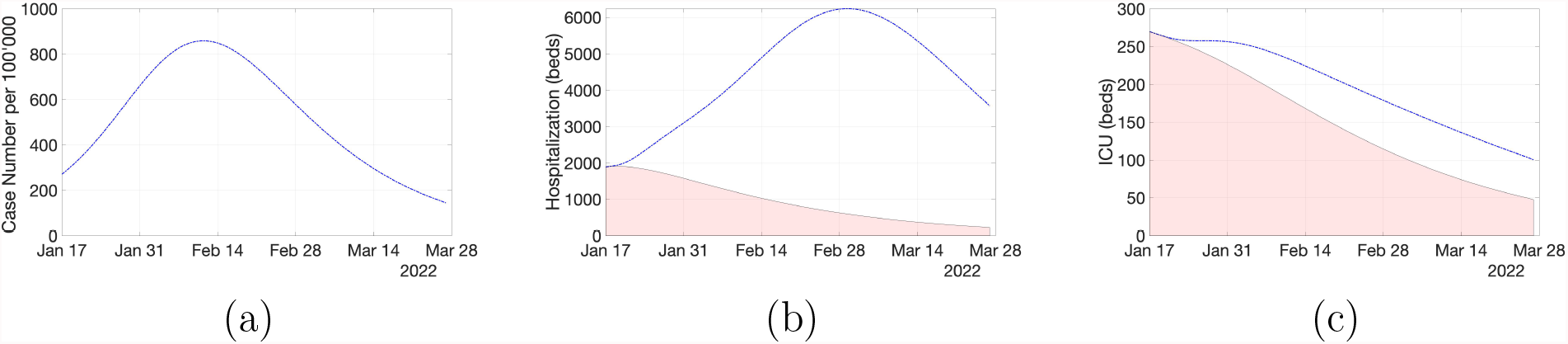
Projection of Scenario 1 in Switzerland at ℛ_*e*_ = 1.8 for (a) daily incidence (Omicron cases), hospitalization (general ward) and (c) ICU occupancy. The average recovery time of hospitalized cases (general ward) is considered to be 4.5 days (in contrast to 1.5 days in the base scenario).

**Figure 5:**
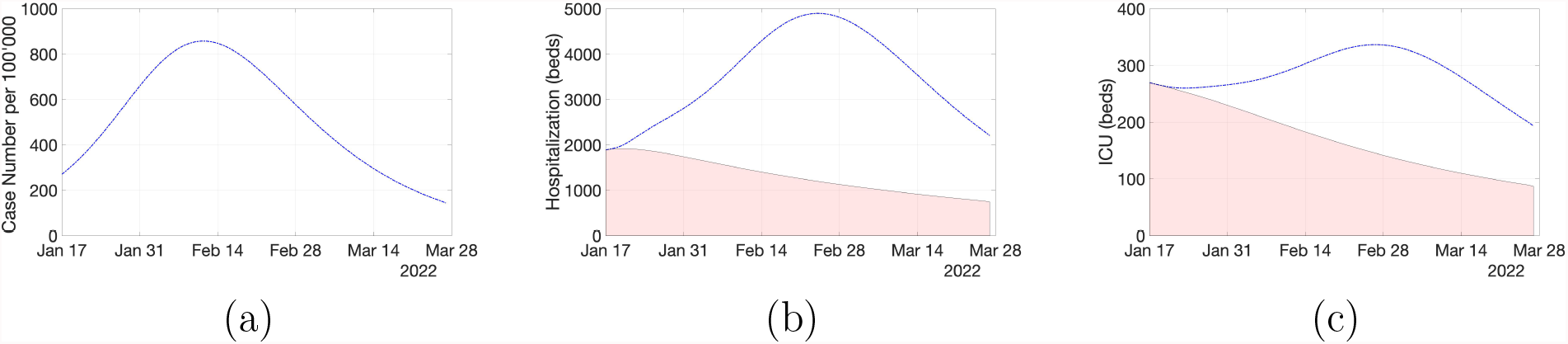
Projection of Scenario 2 in Switzerland at ℛ_*e*_ = 1.8 for (a) daily incidence (Omicron cases), (b) hospitalization (general ward) and (c) ICU occupancy. The severity of Omicron is assumed 50% lower than Delta here.

**Figure 6:**
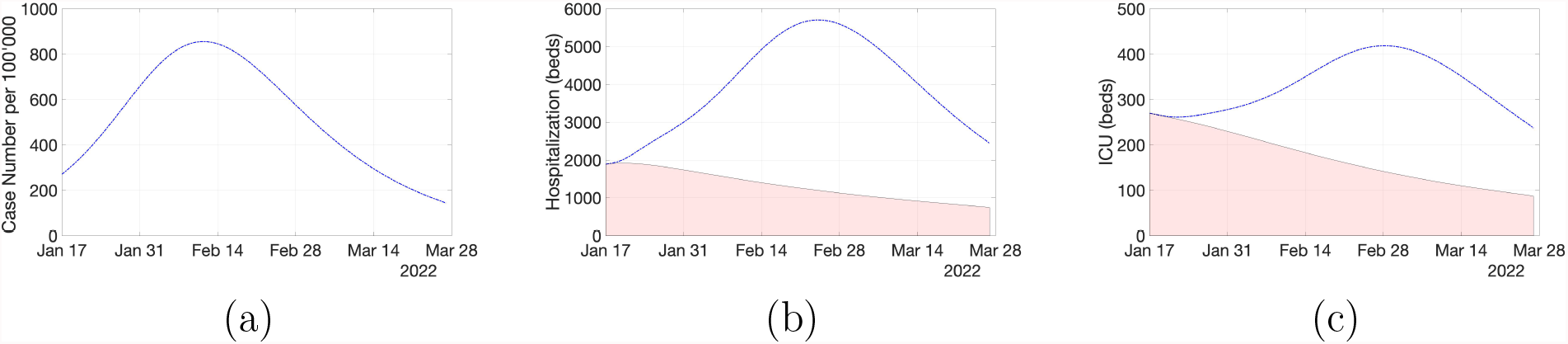
Projection of Scenario 3 in Switzerland at ℛ_*e*_ = 1.8 for (a) daily incidence (Omicron cases), (b) hospitalization (general ward) and (c) ICU occupancy. The severity of Omicron is assumed 40% lower than Delta here and the vaccine efficacy against hospitalization is 5% lower than the base assumption.

**Figure 7:**
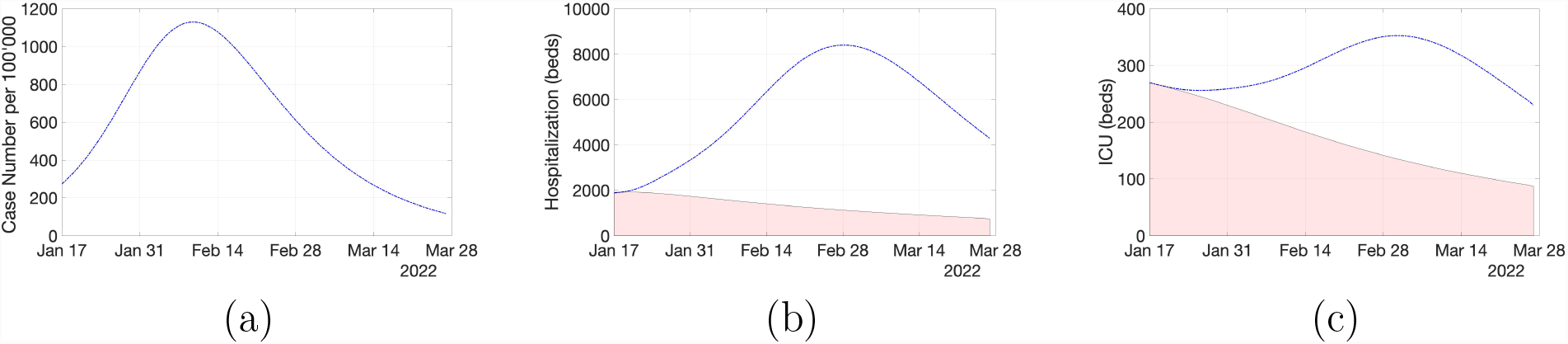
Projection of Scenario 4 in Switzerland at ℛ_*e*_ = 1.8 for (a) daily incidence (Omicron cases), (b) hospitalization (general ward) and (c) ICU occupancy. The severity of Omicron is assumed 30% lower than Delta here and the vaccine efficacies against hospitalization and infection are 5% and 10% lower than the base assumptions, respectively.

**Figure 8:**
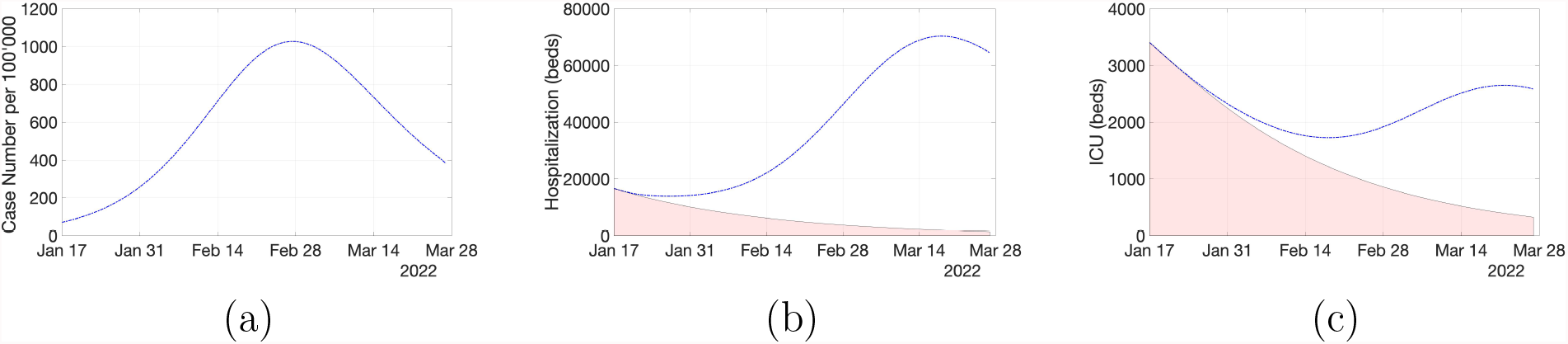
Projection of Scenario 1 in Germany at ℛ_*e*_ = 1.8 for (a) daily incidence (Omicron cases), (b) hospitalization (general ward) and (c) ICU occupancy. The average recovery time of hospitalized cases (general ward) is considered to be 4.5 days (in contrast to 1.5 days in the base scenario).

**Figure 9:**
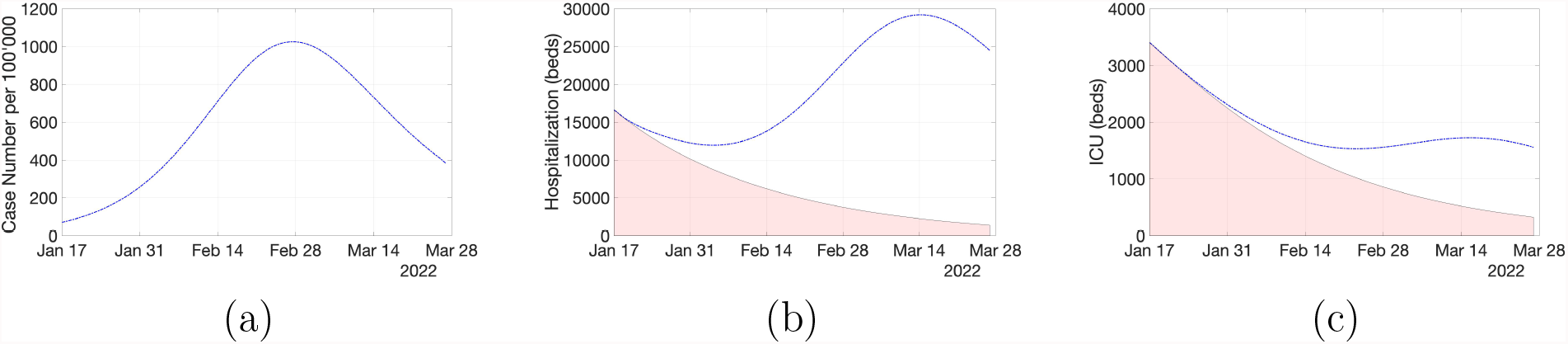
Projection of Scenario 2 in Germany at ℛ_*e*_ = 1.8 for (a) daily incidence (Omicron cases), (b) hospitalization (general ward) and (c) ICU occupancy. The severity of Omicron is assumed 50% lower than Delta here.

**Figure 10:**
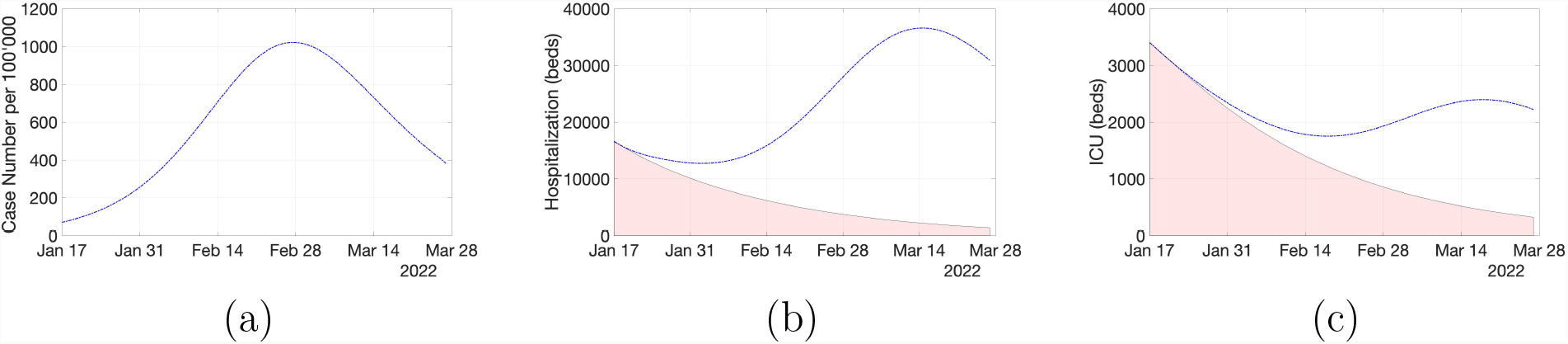
Projection of Scenario 3 in Germany at ℛ_*e*_ = 1.8 for (a) daily incidence (Omicron cases), (b) hospitalization (general ward) and (c) ICU occupancy. The severity of Omicron is assumed 40% lower than Delta here and the vaccine efficacy against hospitalization is 5% lower than the base assumption.

**Figure 11:**
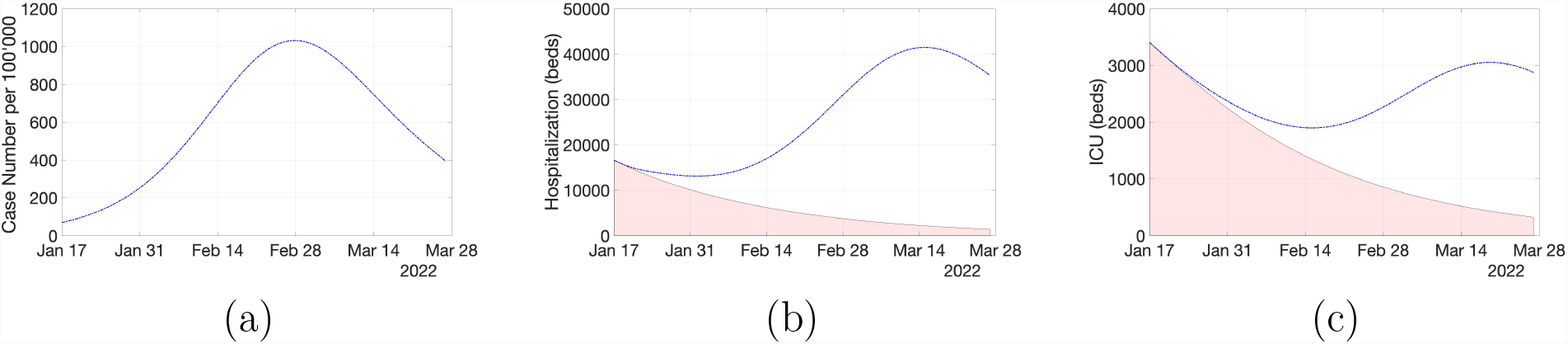
Projection of Scenario 4 in Germany at ℛ_*e*_ = 1.8 for (a) daily incidence (Omicron cases), (b) hospitalization (general ward) and (c) ICU occupancy. The severity of Omicron is assumed 30% lower than Delta here and the vaccine efficacies against hospitalization and infection are 5% and 10% lower than the base assumptions, respectively.

Next, we investigated the effect of age-mixing patterns on the evolution of the pandemic. Figure 12 shows the adopted contact matrices for Switzerland and Germany (in the absence of social-distancing) [21]. It is clear that the contacts are estimated to be significantly more heterogeneous in the case of Switzerland. To evaluate the impact of different age-mixing patterns of the two countries on our results, we study the base scenarios but with swapped contact matrices.

**Figure 12:**
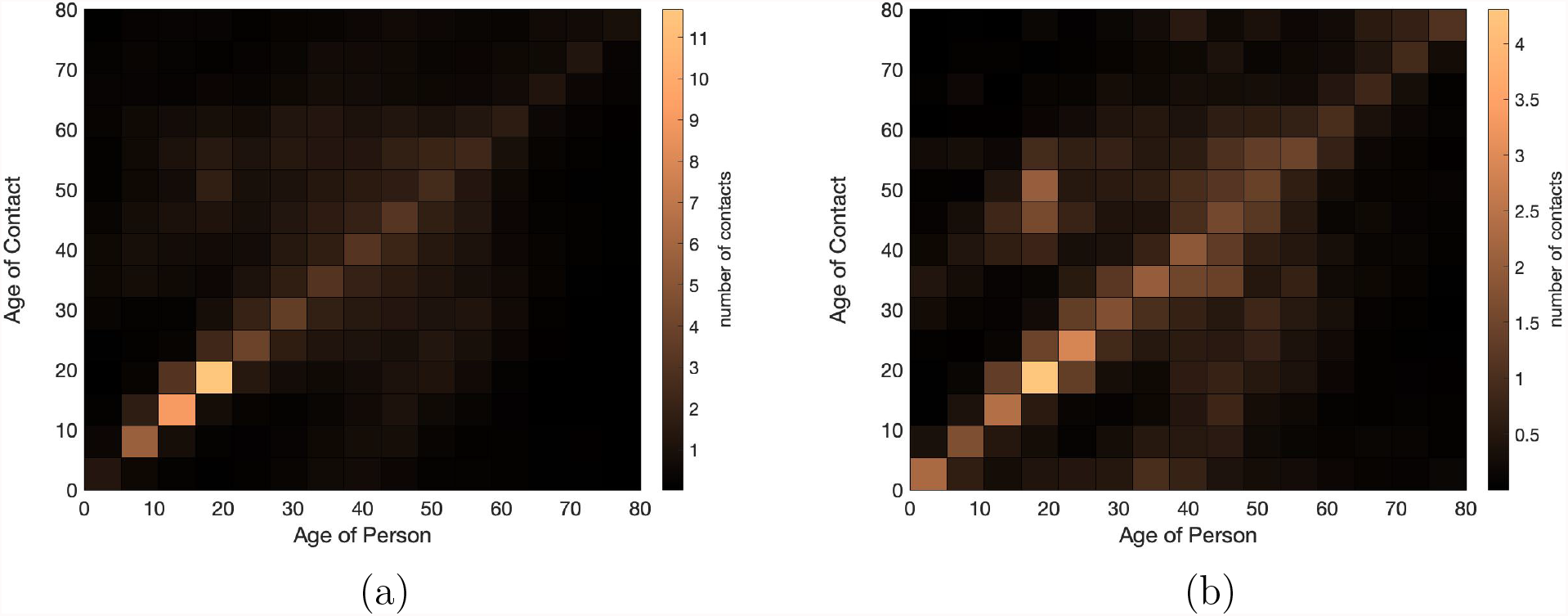
Contact matrices by age for (a) Switzerland and (b) Germany [21].

Figure 13 shows the modeled contact scenarios. As expected, by reducing the heterogeneity between the age-mixing contacts, we observe a higher increase in the case numbers in Switzerland, for the same reproductive number. Conversely, the effect is reversed in Germany where a less uniform contact matrix leads to a lower epidemic burden, for the same reproductive number.

**Figure 13:**
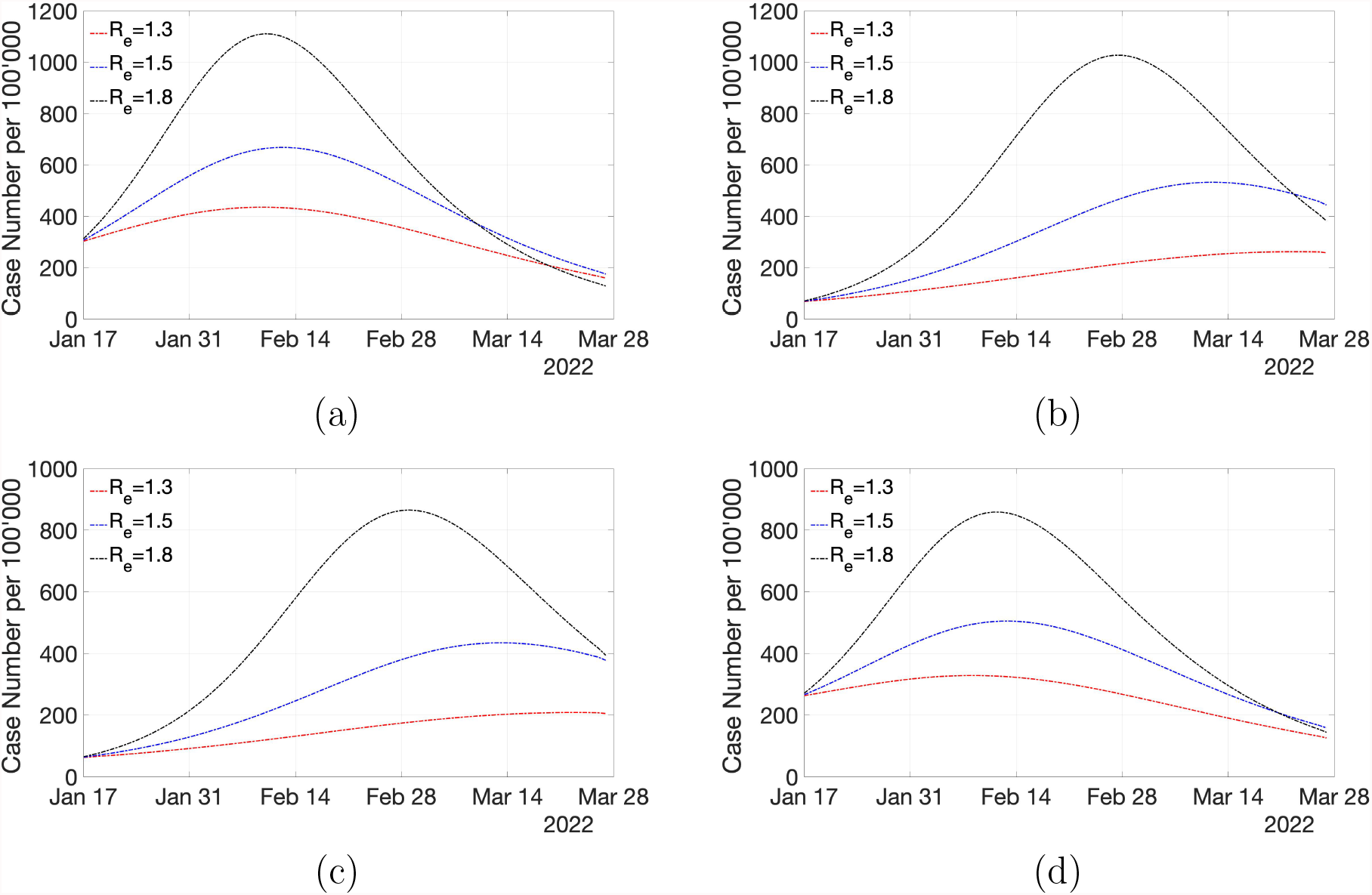
Effect of age-mixing contact matrix on the epidemic wave (a) scenario projection of Switzerland with contact matrix of Germany, (b) scenario projection of Germany (original contact matrix), scenario projection of Germany with contact matrix of Switzerland and (d) scenario projection of Switzerland (original contact matrix).

Overall our sensitivity investigation further supports the main result that the current Omicron wave may not impose a significant threat on the healthcare system of the considered populations. Nevertheless the constant stress on the healthcare demand and possible shortage of staff due to the high infection rate across the population should be considered in evaluating the risk of the current Omicron wave.

## Computation and Datasets

The dynamic model was implemented with MATLAB and the Statistics Toolbox Release 2020b. The vaccine efficacies were implemented with R. The codes are available upon request from the corresponding author. All datasets used in this study are publicly available. The current epidemiological state of Switzerland and Germany are set according to the official data of Federal Office of Public Health (FOPH) and the Robert-Koch-Institute (RKI), respectively. We used contact matrices estimated by [21].

## Data Availability

All datasets used in this study are publicly available. The codes are available upon request from the corresponding author.

## Acknowledgement

This work has been realized as a sub-project of B-FAST (Bundesweites Forschungsnetzwerk Angewandte Surveillance und Testung) for the joint project National Research Network of University Medicine on COVID-19, funded by the Federal Ministry of Education and Research (BMBF - FKZ 01KX2021).

## References

[1] Uk health security agency. sars-cov-2 variants of concern and variants under investigation in england, Technical briefing 312021, (2022).

[2] F. Abdullah, J. Myers, D. Basu, G. Tintinger, V. Ueckermann, M. Mathebula, R. Ramlall, S. Spoor, T. de Villiers, Z. Van der Walt, et al., Decreased severity of disease during the first global omicron variant covid-19 outbreak in a large hospital in tshwane, south africa, International Journal of Infectious Diseases, (2021).

[3] C. L. Althaus, S. Baggio, M. L. Reichmuth, E. B. Hodcroft, J. Riou, R. A. Neher, F. Jacquerioz, H. Spechbach, J. Salamun, P. Vetter, et al., A tale of two variants: Spread of sars-cov-2 variants alpha in geneva, switzerland, and beta in south africa, medRxiv, (2021).

[4] F. Balabdaoui and D. Mohr, Age-stratified discrete compartment model of the covid-19 epidemic with application to switzerland, Scientific reports, 10 (2020), pp. 1–12.

[5] R. C. Barnard, N. G. Davies, C. A. Pearson, M. Jit, and W. John, Modelling the potential consequences of the omicron sars-cov-2 variant in england, 2021.

[6] O. Diekmann, J. Heesterbeek, and M. G. Roberts, The construction of next-generation matrices for compartmental epidemic models, Journal of the Royal Society Interface, 7 (2010), pp. 873–885.

[7] N. Ferguson, D. Laydon, G. Nedjati Gilani, N. Imai, K. Ainslie, M. Baguelin, S. Bhatia, A. Boonyasiri, Z. Cucunuba Perez, G. Cuomo-Dannenburg, et al., Report 9: Impact of non-pharmaceutical interventions (npis) to reduce covid19 mortality and healthcare demand, (2020).

[8] L. Fumanelli, M. Ajelli, P. Manfredi, A. Vespignani, and S. Merler, Inferring the structure of social contacts from demographic data in the analysis of infectious diseases spread, (2012).

[9] S. Funk, J. K. Knapp, E. Lebo, S. E. Reef, A. J. Dabbagh, K. Kretsinger, M. Jit, W. J. Edmunds, and P. M. Strebel, Combining serological and contact data to derive target immunity levels for achieving and maintaining measles elimination, BMC medicine, 17 (2019), pp. 1–12.

[10] H. Gorji, M. Arnoldini, D. F. Jenny, W.-D. Hardt, and P. Jenny, Smart investment of virus rna testing resources to enhance covid-19 mitigation, Plos one, 16 (2021), p. e0259018.

[11] X. He, E. H. Lau, P. Wu, X. Deng, J. Wang, X. Hao, Y. C. Lau, J. Y. Wong, Y. Guan, X. Tan, et al., Temporal dynamics in viral shedding and transmissibility of covid-19, Nature medicine, 26 (2020), pp. 672–675.

[12] L. Jansen, Investigation of a sars-cov-2 b. 1.1. 529 (omicron) variant cluster—nebraska, november– december 2021, MMWR. Morbidity and mortality weekly report, 70 (2021).

[13] D. S. Khoury, M. Steain, J. Triccas, A. Sigal, M. P. Davenport, and D. Cromer, Analysis: A meta-analysis of early results to predict vaccine efficacy against omicron, medRxiv, (2021).

[14] D.-K. Kipourou, C. Leyrat, N. Alsheridah, S. Almazeedi, S. Al-Youha, M. H. Jamal, M. Al-Haddad, S. Al-Sabah, B. Rachet, and A. Belot, Probabilities of icu admission and hospital discharge according to patient characteristics in the designated covid-19 hospital of kuwait, BMC public health, 21 (2021), pp. 1–11.

[15] J. A. Lewnard, V. X. Hong, M. M. Patel, R. Kahn, M. Lipsitch, and S. Y. Tartof, Clinical outcomes among patients infected with omicron (b. 1.1. 529) sars-cov-2 variant in southern california, medRxiv, (2022).

[16] Y. Liu and J. Rocklöv, The reproductive number of the delta variant of sars-cov-2 is far higher compared to the ancestral sars-cov-2 virus, Journal of travel medicine, (2021).

[17] S. Madhi, G. Kwatra, J. E. Myers, W. Jassat, N. Dhar, C. K. Mukendi, A. Nana, L. Blumberg, R. Welch, N. Ngorima-Mabhena, et al., South african population immunity and severe covid-19 with omicron variant, medRxiv, (2021).

[18] S. Mayor, Covid-19: Warning over transmission risk as self-isolation is cut to five days in england, 2022.

[19] P. Nordström, M. Ballin, and A. Nordström, Effectiveness of covid-19 vaccination against risk of symptomatic infection, hospitalization, and death up to 9 months: A swedish total-population cohort study, Hospitalization, and Death Up to, 9 (2021).

[20] S. Paul and E. Lorin, Distribution of incubation periods of covid-19 in the canadian context, Scientific Reports, 11 (2021), pp. 1–9.

[21] K. Prem, A. R. Cook, and M. Jit, Projecting social contact matrices in 152 countries using contact surveys and demographic data, PLoS computational biology, 13 (2017), p. e1005697.

[22] J.-D. Van Wees, M. van der Kuip, S. Osinga, B. Keijser, D. van Westerloo, M. Hanegraaf, M. Pluymaekers, O. Leeuwenburgh, L. Brunner, and M. T. van Furth, Sir model for assessing the impact of the advent of omicron and mitigating measures on infection pressure and hospitalization needs, MedRxIv, (2021).

[23] L. Veneti, B. V. Salamanca, E. Seppälä, J. Starrfelt, M. L. Storm, K. Bragstad, O. Hungnes, H. Bøås, R. Kvåle, L. Vold, et al., No difference in risk of hospitalisation between reported cases of the sars-cov-2 delta variant and alpha variant in norway, International Journal of Infectious Diseases, (2021).

[24] R. Viana, S. Moyo, D. G. Amoako, H. Tegally, C. Scheepers, R. J. Lessells, J. Giandhari, N. Wolter, J. Everatt, A. Rambaut, et al., Rapid epidemic expansion of the sars-cov-2 omicron variant in southern africa, medRxiv, (2021).

[25] L. Wang, N. A. Berger, D. C. Kaelber, P. B. Davis, N. D. Volkow, and R. Xu, Comparison of outcomes from covid infection in pediatric and adult patients before and after the emergence of omicron, medRxiv, (2022), pp. 2021–12.

[26] N. Wolter, W. Jassat, S. Walaza, R. Welch, H. Moultrie, M. Groome, D. G. Amoako, J. Everatt, J. N. Bhiman, C. Scheepers, et al., Early assessment of the clinical severity of the sars-cov-2 omicron variant in south africa, medRxiv, (2021).

[27] F. Zhou, T. Yu, R. Du, G. Fan, Y. Liu, Z. Liu, J. Xiang, Y. Wang, B. Song, X. Gu, et al., Clinical course and risk factors for mortality of adult inpatients with covid-19 in wuhan, china: a retrospective cohort study, The lancet, 395 (2020), pp. 1054–1062.

